# Widening geographic range of Rift Valley fever disease clusters associated with climate change in East Africa

**DOI:** 10.1101/2024.05.17.24307534

**Authors:** Silvia Situma, Luke Nyakarahuka, Evans Omondi, Marianne Mureithi, Marshal Mweu, Matthew Muturi, Athman Mwatondo, Jeanette Dawa, Limbaso Konongoi, Samoel Khamadi, Erin Clancey, Eric Lofgren, Eric Osoro, Isaac Ngere, Robert F. Breiman, Barnabas Bakamutumaho, Allan Muruta, John Gachohi, Samuel O. Oyola, M. Kariuki Njenga, Deepti Singh

## Abstract

**Background:** Recent epidemiology of Rift Valley fever (RVF) disease in Africa suggests growing frequency and expanding geographic range of small disease clusters in regions that previously had not reported the disease. We investigated factors associated with the phenomenon by characterizing recent RVF disease events in East Africa.

**Methods:** Data on 100 disease events (2008 – 2022) from Kenya, Uganda, and Tanzania were obtained from public databases and institutions, and modeled against possible geo-ecological risk factors of occurrence including altitude, soil type, rainfall/precipitation, temperature, normalized difference vegetation index (NDVI), livestock production system, land-use change, and long-term climatic variations. Decadal climatic variations between 1980-2022 were evaluated for association with the changing disease pattern.

**Results:** Of 100 events, 91% were small RVF clusters with a median of one human (IQR, 1-3) and 3 livestock cases (IQR, 2-7). These clusters exhibited minimal human mortality (IQR 0-1), and occurred primarily in highlands (67%), with 35% reported in areas that had never reported RVF disease. Multivariate regression analysis of geo-ecological variables showed a positive correlation between occurrence and increasing temperature and rainfall. A 1oC increase in temperature and 1-unit increase in NDVI, 1-3 months prior were associated with increased RVF incidence rate ratios (IRR) of 1.20 (95% CI 1.1,1.2) and 9.88 (95% CI 0.85, 119.52), respectively. Long-term climatic trends showed significant decadal increase in annual mean temperature (0.12 to 0.3oC/decade, P<0.05), associated with decreasing rainfall in arid and semi-arid lowlands but increasing rainfall trends in highlands (P<0.05). These hotter and wetter highlands showed increasing frequency of RVF clusters, accounting for 76% and 43% in Uganda and Kenya, respectively.

**Conclusion:** These findings demonstrate the changing epidemiology of RVF disease. The widening geographic range of disease is associated with climatic variations, with the likely impact of wider dispersal of virus to new areas of endemicity and future epidemics.

**Key questions:** *What is already known on this topic?:* Rift Valley fever is recognized for its association with heavy rainfall, flooding, and El Niño rains in the East African region. A growing body of recent studies has highlighted a shifting landscape of the disease, marked by an expanding geographic range and an increasing number of small RVF clusters.

*What this study adds:* This study challenges previous beliefs about RVF, revealing that it predominantly occurs in small clusters rather than large outbreaks, and its association with El Niño is not as pronounced as previously thought. Over 65% of these clusters are concentrated in the highlands of Kenya and Uganda, with 35% occurring in previously unaffected regions, accompanied by an increase in temperature and total rainfall between 1980 and 2022, along with a rise in the annual number of rainy days. Notably, the observed rainfall increases are particularly significant during the short-rains season (October-December), aligning with a secondary peak in RVF incidence. In contrast, the lowlands of East Africa, where typical RVF epidemics occur, display smaller and more varied trends in annual rainfall.

*How this study might affect research, practice, or policy:* The worldwide consequence of the expanding RVF cluster is the broader dispersion of the virus, leading to the establishment of new regions with virus endemicity. This escalation heightens the risk of more extensive extreme-weather-associated RVF epidemics in the future. Global public health institutions must persist in developing preparedness and response strategies for such scenarios. This involves the creation and approval of human RVF vaccines and therapeutics, coupled with a rapid distribution plan through regional banks.

## INTRODUCTION

Rift Valley fever (RVF) virus, a mosquito-borne phlebovirus, is known for extreme weather-associated large epidemics characterized by abortions and perinatal mortalities in livestock; and fever, jaundice, encephalitis, retinitis, and hemorrhagic syndrome in humans. [1] [2] First isolated in Kenya in 1931, the virus caused sporadic localized clusters of RVF disease during the rainy season until 1951 when a larger epidemic spread the virus to approximately 33% of the country. [3] Since the 1960s Kenya has reported >10 major RVF epidemics associated with El Niño/Southern Oscillation (ENSO) heavy rainfall, including the two most extensive regional epidemics (Somalia, Kenya, Tanzania) in 1997-98 and 2006-07 that attracted the involvement of international partners including World Health Organization as public health events of international concern. [4] From the 1970s, periodic major epidemics have been reported in the eastern (Kenya, Somalia, Tanzania), southern (Zimbabwe, South Africa, Madagascar, Mayotte), northern (Egypt, Tunisia), and western (Senegal, Mali, Niger, Mauritania) regions of Africa, almost all associated with flooding conditions. [4] In 2000, the virus crossed over to the Middle East, through livestock trade, causing a major epidemic in Saudi Arabia and Yemen that resulted in over 3500 confirmed human cases and >200 fatalities. [5] [6]

Recurrent RVF epidemics occur in certain ecosystems where the virus is endemic, following the convergence of several factors including the presence of mosquito vectors, adequate density of naïve livestock, and extreme weather conditions that bring excessive flooding to trigger large-scale vector populations. [7] In East Africa, such permissive ecologies, which were responsible for up to 90% of human cases during epidemics, were characterized by low-elevation plains (<1000 m above sea level) with slow-draining clay soil, and dense bushes or acacia vegetations. [8] RVF epidemics can also occur in new areas, triggered by massive livestock movement as observed in Africa to Middle East livestock trade during annual Muslim Hajj, or political unrest resulting in largescale displacement of communities. [9] [10]

Once the virus is introduced in a country or region, it becomes endemic through maintenance by *Aedes* mosquitoes via transovarial transmission. [11] [12] The virus can also be maintained through continuous low-level cycling in livestock, herbivore wildlife, and humans and mosquitoes. [13] [14] During such cryptic cycles, a small number of infected mosquitoes routinely emerges to infect animals and humans; however, this cycling does not reach the epidemic threshold, either because the infected vector population is small and rapidly dies off, or a second vector responsible for the horizontal transmissions that occur in epidemics is not present in sufficient numbers. [15] [16]

There is a growing global concern that changing climate conditions and increasing extreme weather events are changing the landscape of infectious diseases by expanding permissive ecologies for pathogens, dispersing pathogens to new regions, and bringing humans closer to pathogens through land use changes. [17] Recent RVF studies pointed to an expanding geographic range and scope of the disease, likely associated with increasing rainfall intensity and or warming temperatures that perhaps broaden ecological areas supportive of virus maintenance. [18] For example, a modeling study conducted in 2017-2018 in the greater horn of Africa (Ethiopia, Kenya, Somalia, Uganda, and Tanzania) using data from epidemics, small RVF clusters (≤20 human cases), and seroprevalence findings identified a broader set of risk factors of RVF occurrence, including new soil types (nitisols, vertisols), both low and high elevation areas, highly variable temperatures during the driest months. [13] In addition, Uganda, which was never involved in severe RVF epidemic occurring in East Africa, started reporting an increasing number of RVF outbreaks in 2016, and significant disease seroprevalence primarily in the highlands of western and central regions of the country. [19] [20] To gain an understanding of the changing RVF epidemiology, we investigated the geo-ecological and climatic factors associated with the spatiotemporal occurrence of small RVF clusters in Kenya, Uganda, and Tanzania between 2008-2022.

## METHODS

### Study area

The study was conducted in the East Africa region (Kenya, Uganda, and Tanzania) which is characterized by a diverse ecosystem ranging from agriculturally productive highlands to semiarid and arid lands inhabited by nomadic pastoralists. [21] Regional climatic conditions are heterogeneous, with Kenya, Uganda, and northeastern Tanzania experiencing bimodal rainfall season with long rains occurring from March to May and short rains from October to December. [22] In contrast, central and southern Tanzania experience unimodal rainfall from November to May. [22] Most of the region experiences substantial rainfall levels in at least one season except the northern and eastern parts of Kenya. [22] Temperatures in most areas range from a moderate 15°C – 25°C except in coastal belts characterized by humidity and higher temperatures. [22] In the arid and semiarid regions of northern Kenya and Uganda, dry and hotter conditions prevail. [22]

### Collection of RVF disease data

For this study, an RVF disease event was defined as a report of suspected RVF cases in either humans or livestock, confirmed by the detection of viral RNA, or antiviral IgM antibodies within a specific location and within a duration of 21 days. The study used presence-only data for disease events and classified each event as either a small cluster if reporting ≤20 confirmed cases (human and animal), or an outbreak characterized by >20 confirmed human and animal cases. This classification was based on our review of RVF disease events reported during inter-epidemic periods in Africa and designed to exclude large outbreaks typically associated with extreme weather conditions and flooding from the analysis.

We collected RVF disease data spanning from 2008 to 2022 in Kenya and Tanzania, and from 2016 to 2022 in Uganda. The data were sourced from Ministries of Health and Agriculture databases, national research institutions, and both published and unpublished reports.[3] [19] [20] [23] - [38] The search strategy for disease events employed precise search strings incorporating keywords like “Rift Valley Fever” “RVF,” “East Africa,” “Kenya,” “Tanzania,” “Uganda,” “IgM,” and “PCR.” We utilized Boolean operators “AND” and “OR” to logically combine these terms, focusing on identifying relevant literature on RVF outbreaks/cases in East Africa. For each RVF disease event, exact geo-coordinates, or geo-coordinates of the area district centroid, start and end date, animal and human involvement, human fatalities, previous history of RVF occurrence, human and livestock population and densities, land and climatic factors were recorded.

### Collection of risk factor data

Data on risk factors such as livestock production systems, land use changes, excessive rainfall, and the presence of significant water accumulation were extracted from disease surveillance reports, online databases, and peer-reviewed publications. [3] [30] [32] [35] Information on proximity to wildlife conservation, soil types, and elevation level was extracted from MODIS satellite images [39] and Food and Agriculture Organization (FAO) databases. [40] Soil types were categorized based on their physical and chemical properties using the FAO scheme. [40] Furthermore, data on animal and human related factors, including predominant livestock species, livestock, and human population densities were gathered. [41] [42] [43] [44] [45] Population and density data for both humans and livestock covering the period from 2008 to 2022 were obtained from the National census data of Kenya, [41] Uganda, [42] Tanzania, [43] [44] and the World Population databases. [45]

### Short-term and long-term climate data

Short-term climatic variables included rainfall, temperature, humidity, and normalized differential vegetation index (NDVI) data from 3 years and 3 months prior to disease event were obtained from Climate Hazards Group InfraRed Precipitation with Station data (CHIRPS), [46] Climate Hazards Group InfraRed Temperature with Station data (CHIRTS), [47] and FAO databases. [40] The study further examined long-term trends in daily mean temperature and precipitation metrics over the past four decades (1981-2022), utilizing high resolution data of 0.05 degrees, specifically daily precipitation data and monthly temperature data from the (CHIRPS) [46] and Climate Research Unit gridded time-series (CRU TS v 4.07).[48] [49] These data sets were employed to quantify seasonal and annual total rainfall, determine the annual number of dry days each (days with precipitation ≤1mm/day) and calculate annual and seasonal mean temperatures.

### Data analysis

For risk factors analysis, we assessed the impact of 45 variables associated with demographic, climatic, and anthropogenic predictors, soil types, altitude, distance to wildlife conservation, predominant livestock species, livestock production system, monthly (up to 3 months prior), and annual (up to 3 years prior) NDVI, temperature, rainfall, and humidity (Supplement Table I). [50] [51] Using total number of RVF cases (both human and livestock) as the dependent variable, we conducted bivariate negative binomial regression to model the number of RVF cases (livestock and human combined) with an offset term (total host density minus number of hosts per km^2^) and reported as incident rate ratios (IRR). For continuous variables with missing values, imputation was performed using a single imputation method.[52] To address collinearity, we conducted pairwise correlations among predictor variables, identifying high correlations among several of them particularly those describing rainfall, temperature, and humidity, as well as intercorrelations between temperature, rainfall, humidity, and NDVI. To mitigate multicollinearity, the model included only first-month rainfall in the outbreak year and 3rd-month temperature in the outbreak year. Certain variables that had missing values and those not statistically significant in bivariate analysis were dropped. A total of 6 predictor variables with a significance level of p ≤ 0.05 (country, temperature, two NDVI variables), and those historically known to be associated with RVFV activity (humidity and soil type), were included in the final multivariate regression model. This model was constructed using a forward variable selection approach with a p-value threshold of ≤0.05. The final model was refined to include variables without significant collinearity, as indicated by a variance inflation factor (VIF) ≤ 5 and the lowest Akaike Information Criterion (AIC).

For long-term climate data analysis, daily gridded precipitation data from CHIRPS and monthly gridded temperature data from the Climate Research Unit gridded timeseries (CRU TS v 4.07) were utilized for trend analysis. Spatial patterns of long-term trends were visualized using Python packages: namely *matplotlib, geocat, xarray*, and *scipy*. Linear trends and their associated p-values were determined through the linear least-squares regression function (linregress) within the *scipy* package. The significance of trends was assessed using the Wald test, and maps were employed to display significance levels for p-values<0.1. It is important to highlight that all climate data analyses and visualizations were performed using the specified Python packages, and the significance threshold for p-values was set at 0.05. We also plotted temporal climate trends against the number of small RVF clusters.

### Ethical approval

Data access, usage and overall study approach were reviewed and approved by Kenyatta National Hospital - University of Nairobi ethical review committee (KNH-UoN ERC P810/10/2022) for Kenya. Uganda data access and use were approved by the Uganda Ministry of Health, while all Tanzania data were obtained from public databases and publications.

## RESULTS

### Characteristics and spatial distribution of small RVF clusters

We collected data on 100 RVF disease events in East Africa from 2008 through 2022. Of the 100 events, 44 were reported in Uganda, 41 in Kenya, and 15 in Tanzania. Among these, 91 were classified as small clusters, characterized by a median of 3 human cases (IQR, 1-3) and 3 livestock cases (IQR, 1-7), with minimal human mortality (IQR 0-1) as detailed in Table 1. Thirty five percent (35%) of the clusters occurred in regions that had never reported RVF cases before, spanning a broader range of altitudes as low as 77m to as high as 2867m above sea level, with a median altitude of 1224m. In Uganda, 93% of the clusters were situated in altitudes ≥1000m, with 48% (17/35) in Kenya, and 25% (3/12) in Tanzania (Table 1, Figure 1). Geographically, 43% (15/35) of the small RVF clusters in Kenya were concentrated in the Southwestern highlands, while a 75 % (33/44) in Uganda occurred in the Southcentral highlands (Figure 1). The southcentral Kenya and southwestern Uganda regions are agriculturally productive highlands, characterized by intensive and semi-intensive livestock production systems. Overall, small clusters occurred across the entire spectrum of livestock production systems, including intensive, semi-intensive, and extensive, whereas RVF epidemics were primarily reported in dry areas with extensive livestock production systems. The majority (65%) of the soils where the small RVF clusters occurred were clayey, specifically nitisols, planosols, and ferralsols soil types, followed by loamy (18%), and sandy soil types (15%).

**Figure 1.**
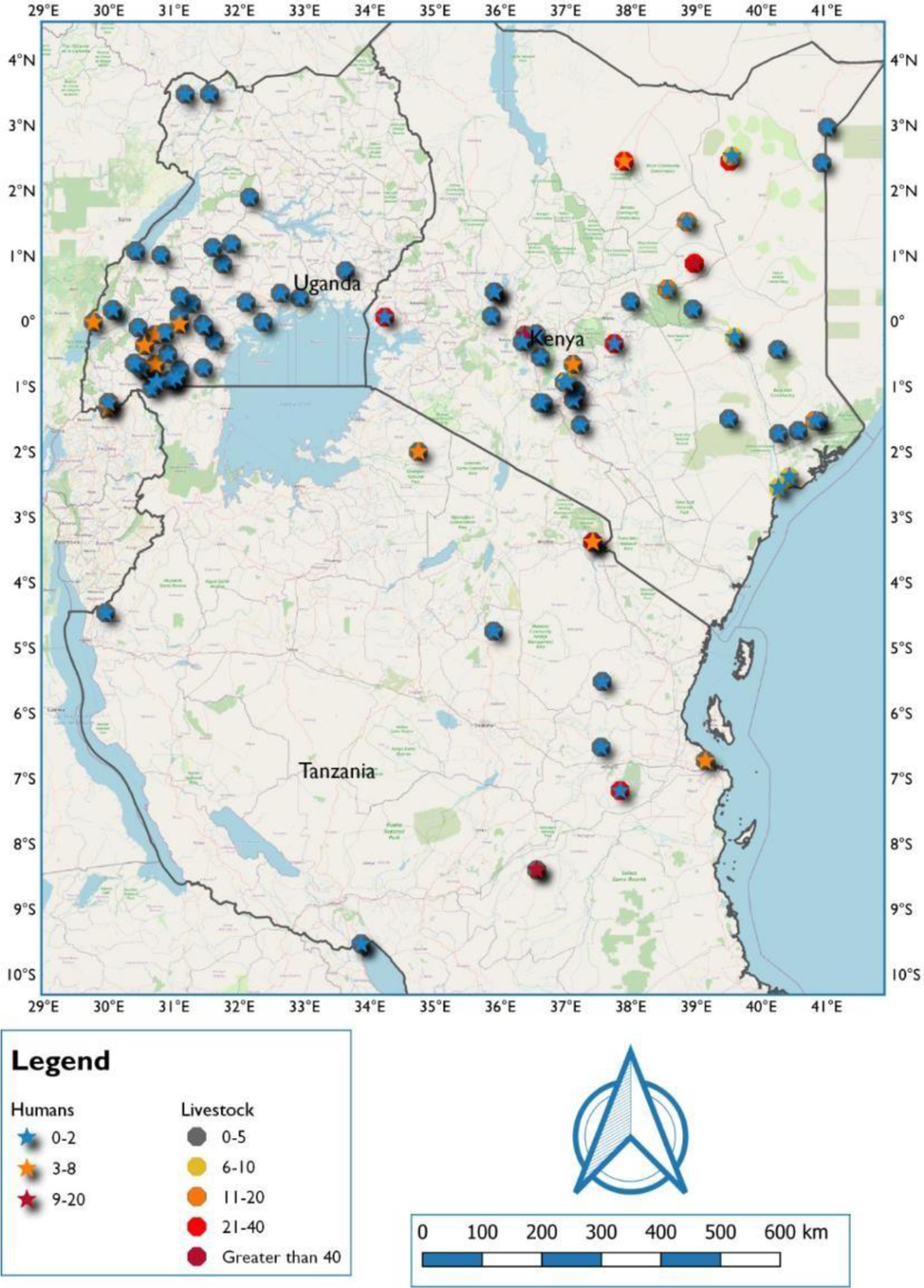
Location where small RVF clusters reported in East Africa between 2008 and 2022

**Table 1:** Characteristics of small RVF clusters in East Africa, 2008-2022, *Note: N= RVF disease events, n= sporadic RVF clusters*.

|  | Kenya | Uganda | Tanzania | Total |
| --- | --- | --- | --- | --- |
| Characteristics | (n=35) | (n= 44) | (n=12) | (n=91) |
| Median no. of human cases (IQR) | 3 (2-3) | 1 (1-2) | 1 (1-6) | 1 (1-3) |
| Median no. of livestock cases (IQR) | 3 (2-7) | 1 (1-1) | 9 (3-14) | 3 (1-7) |
| Median human case mortality | 0 (0) | 1 (0-1) | 0 (0) | 1 (0-1) |
| No. of clusters in new areas | 6 (17%) | 23 (52%) | 3 (25%) | 32(35%) |
| Types of Livestock Production Systems |  |  |  |  |
| Intensive | 14 (40%) | 3 (7%) | 3 (25%) | 20 (22%) |
| semi-intensive | 1 (3%) | 38 (86%) | 2 (17%) | 41 (45%) |
| Extensive | 20 (57%) | 3 (7%) | 7 (58%) | 30 (33%) |
| Soil types |  |  |  |  |
| Clay | 23 (68%) | 24 (67%) | 7 (58%) | 54 (65%) |
| Loam | 8 (23%) | 3 (8%) | 4 (33%) | 15 (18%) |
| Sand | 3 (9%) | 9 (25%) | 1 (8%) | 13 (15%) |
| No. of RVF events with positive NDVI change 1 month prior to cases |  |  |  |  |
| Yes | 20 (59%) | 20 (51%) | 4 (36%) | 43 (51%) |
| No | 14 (41%) | 19 (49%) | 7 (64%) | 41 (49%) |
| Median altitude (m) (IQR) | 743.8 (1429) | 1276 (304) | 491 (769) | 1224 (994) |

*Note: n= sporadic RVF clusters*

A positive NDVI change one month prior to RVF occurrence was observed in 51% of the small clusters. The small RVF clusters were situated in areas with notable land use changes, particularly those leading to significant water accumulation such as rice farming, mining, and irrigation. In areas with water pools, either a result of natural or man-made conditions, (72%) of the small RVF clusters occurred. On average (28%) of the small RVF clusters were in areas with man-made pools resulting from construction of ponds or dams, swamps, irrigation schemes, and excavated mines.

### Seasonality and short-term geo-ecological determinants

When plotted by month of occurrence, the small RVF clusters demonstrated a higher frequency (75/91, 82%) during and immediately after the wet seasons as shown in Figure 2. In bivariate analysis, a history of previous RVF occurrence, the presence of significant water pools, various types of land use changes, both long-term and short-term temperature variations, and increases in NDVI changes were associated with an elevated IRR with p-vales of ≤0.05. (Supplement Table II). Results from the multivariate analysis are presented in Table 2. The multivariate model in Supplement Table III exhibited a higher AIC indicating a poorer fit compared to the model presented in Table 2. Temperature and NDVI demonstrated a significant positive association with the occurrence of small RVF clusters while soil types and elevation were not found to be associated with the changes in IRR. A 1-unit positive change in NDVI a month prior to the diseaseoccurrence, or 1^0^C temperature rise in the 3^rd^ month prior to occurrence were associated with disease IRR of 9.88 (95% CI 0.85, 119.52) and 1.20 (95% CI 1.1,1.2), respectively as shown in Table 2.

**Figure 2.**
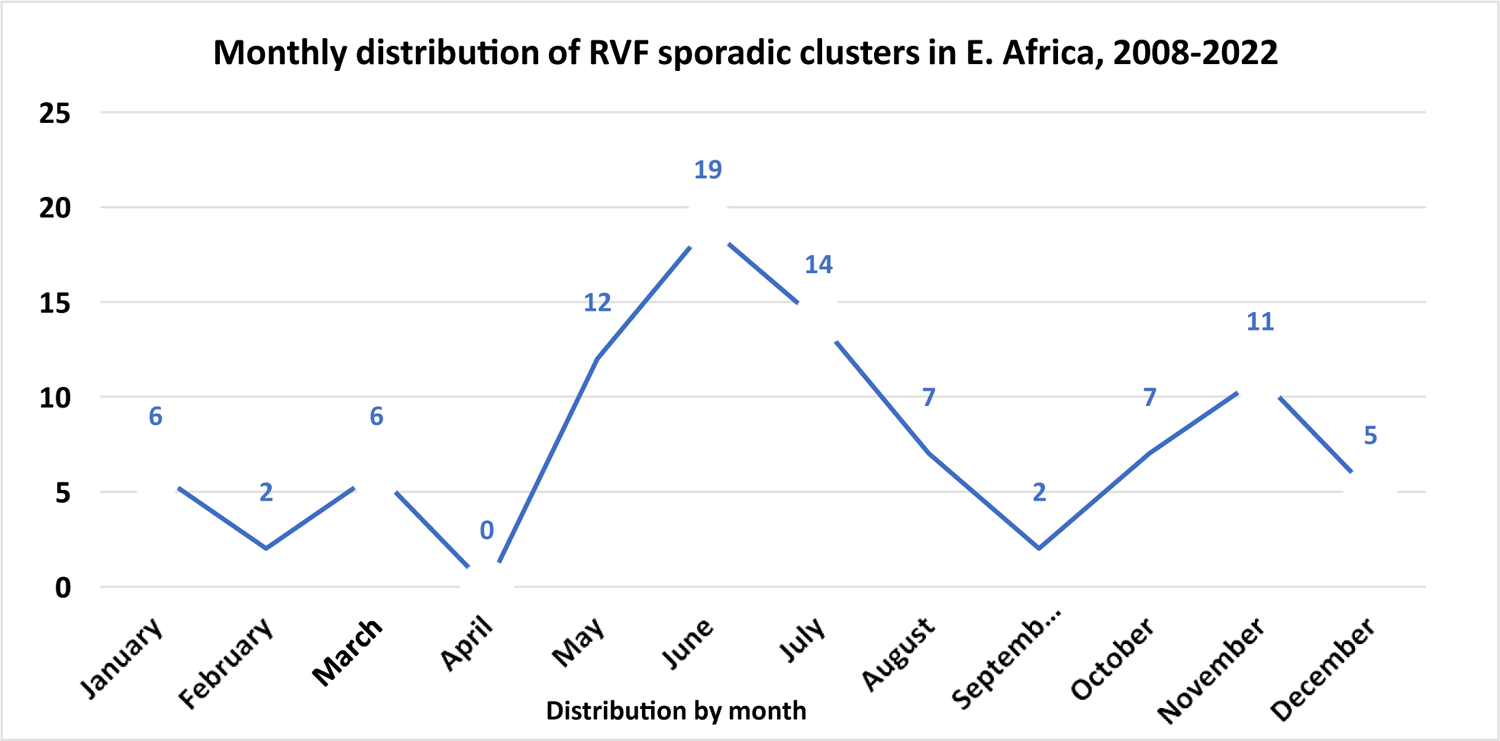
Seasonal distribution of RVF clusters reported in East Africa between 2008 and 2022

**Table 2:** Multivariate analysis of potential drivers of RVF clusters in East Africa, 2008 – 2022.

| Factor | Coefficient | IRR | 95% CI | S.E | p-value |
| --- | --- | --- | --- | --- | --- |
| *Intercept* | -10.53 | - | - | - | - |
| Tanzania | 0.90 | 2.45 | 1.07, 5.92 | 0.42 | 0.03 |
| Uganda | -1.22 | 0.30 | 0.15, 0.57 | 0.32 | <0.001 |
| Temperature (3<sup>rd</sup> month prior) | 0.18 | 1.20 | 1.11, 1.29 | 0.04 | **<0.001** |
| Humidity (1<sup>st</sup> month prior) | -0.01 | 0.99 | 0.95, 1.02 | 0.02 | 0.48 |
| First month prior NDVI | 2.29 | 9.88 | 0.85, 119.52 | 1.05 | **0.03** |
| Positive NDVI change | 0.66 | 1.93 | 1.01, 3.71 | 0.30 | **0.03** |
| Loam soils | -0.20 | 0.82 | 0.39, 1.76 | 0.36 | 0.58 |
| Sandy soils | -0.50 | 0.60 | 0.21, 1.74 | 0.48 | 0.30 |
| Deviance | - | - | **DF** | - | - |
| Null Deviance | 149.52 |  | 75 | - | - |
| Residual Deviance | 76.28 |  | 67 | - | - |
| Dispersion parameter | 1.00 |  | - | - | - |
| AIC | 423.69 |  | - | - | - |
| 2 x log-likelihood | - 403.69 |  | - | - | - |

*Theta: 1.002; Std. Err.: 0.175, Significance codes: 0 '\*\*\*' 0.001 '\*\*' 0.01 '\*' 0.05 '.' 0.1 ' ' 1*
Temperature (3<sup>rd</sup> month prior) refers to the temperature of the 3<sup>rd</sup> month prior to RVF disease event, Humidity (1<sup>st</sup> month prior) refers to humidity of the 1<sup>st</sup> month prior to RVF disease event occurrence, First month prior NDVI refers to NDVI value of the 1<sup>st</sup> month before RVF disease event

### Long-term climatic trends in East Africa, 1981-2022

There is substantial spatial heterogeneity in climatological temperature (Figure 3a,b,c,d) and rainfall patterns (Figure 3,e,f,g,h) across East Africa, partly associated with the topographic complexity of the region. (Note: climatology is calculated as the 30-year average over the current normal period 1991-2020 as defined by the World Meteorological Organization). Annual mean temperatures show substantial warming across the entire region, with larger trends (calculated over 1981-2022) across most of Kenya than in Uganda and Tanzania (Figure 4a). We analyzed the spatial patterns of trends during the long (March - May) and short (March - May) rains season; 82.4% of the RVF cluster occurred during and after these seasons. Temperature trends during the long rains varied between 0.12 (°C/decade) in southwestern Uganda to 0.3 (°C/decade) in northern Kenya (Figure 3b). This warming trend is more widespread and amplified across all three countries during the short rainy season (Figure 3d). Trends in rainfall are mixed, showing increasing wetness in southwestern Uganda, western Kenya, and most of Tanzania but increasing dryness in northwestern Uganda and the expansive low-elevation eastern Kenya region stretching from the costal south to the arid and semi-arid regions north during the long rainy season (Figure 3e,f). In contrast, the short rains season experienced stronger and more widespread wetting trends across Kenya, Uganda, and northwestern Tanzania but strong decreasing trends across southern Tanzania (Figure 3g,h).

**Figure 3.**
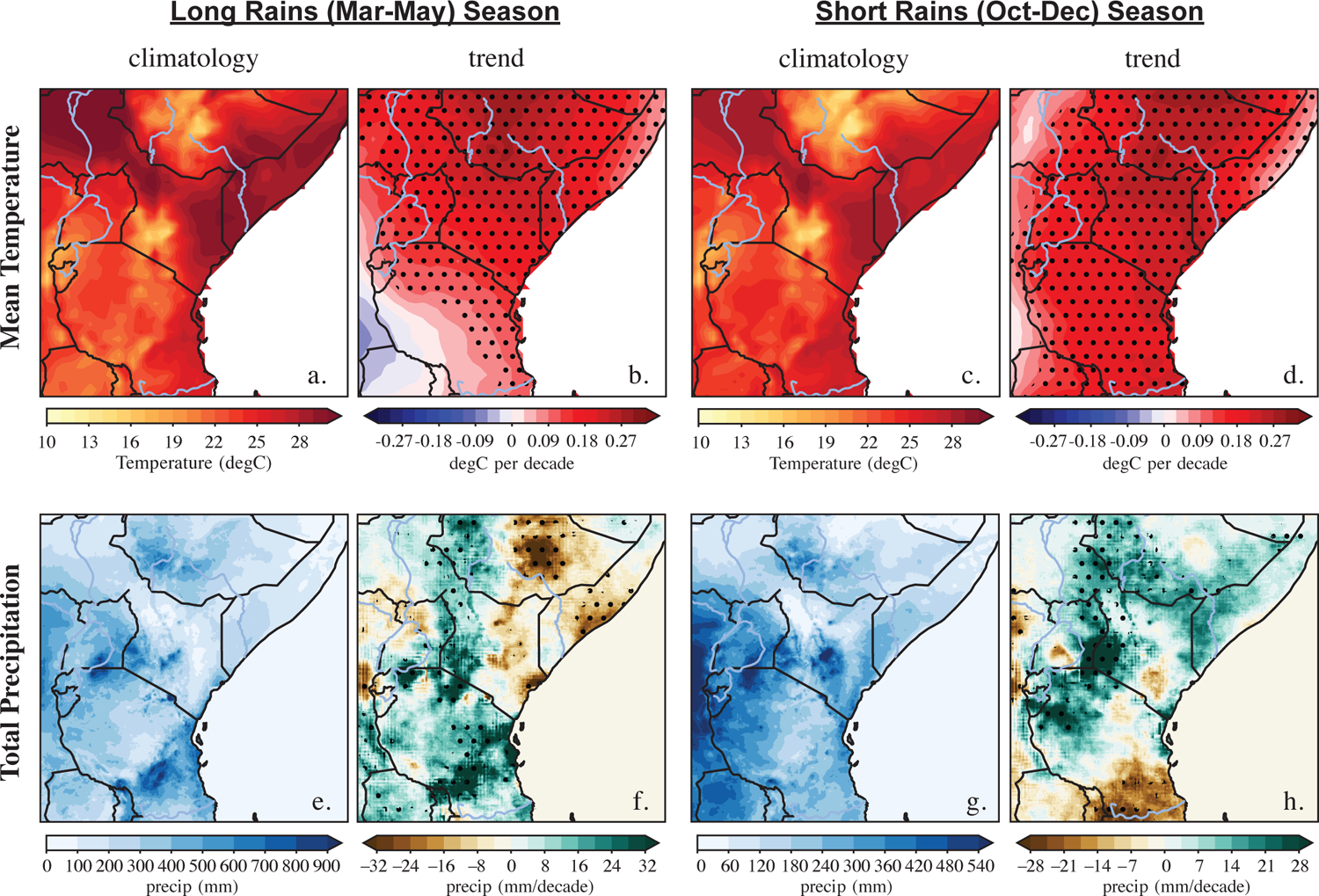
Seasonal temperature and rainfall patterns in Kenya, Uganda, and Tanzania

**Figure 4.**
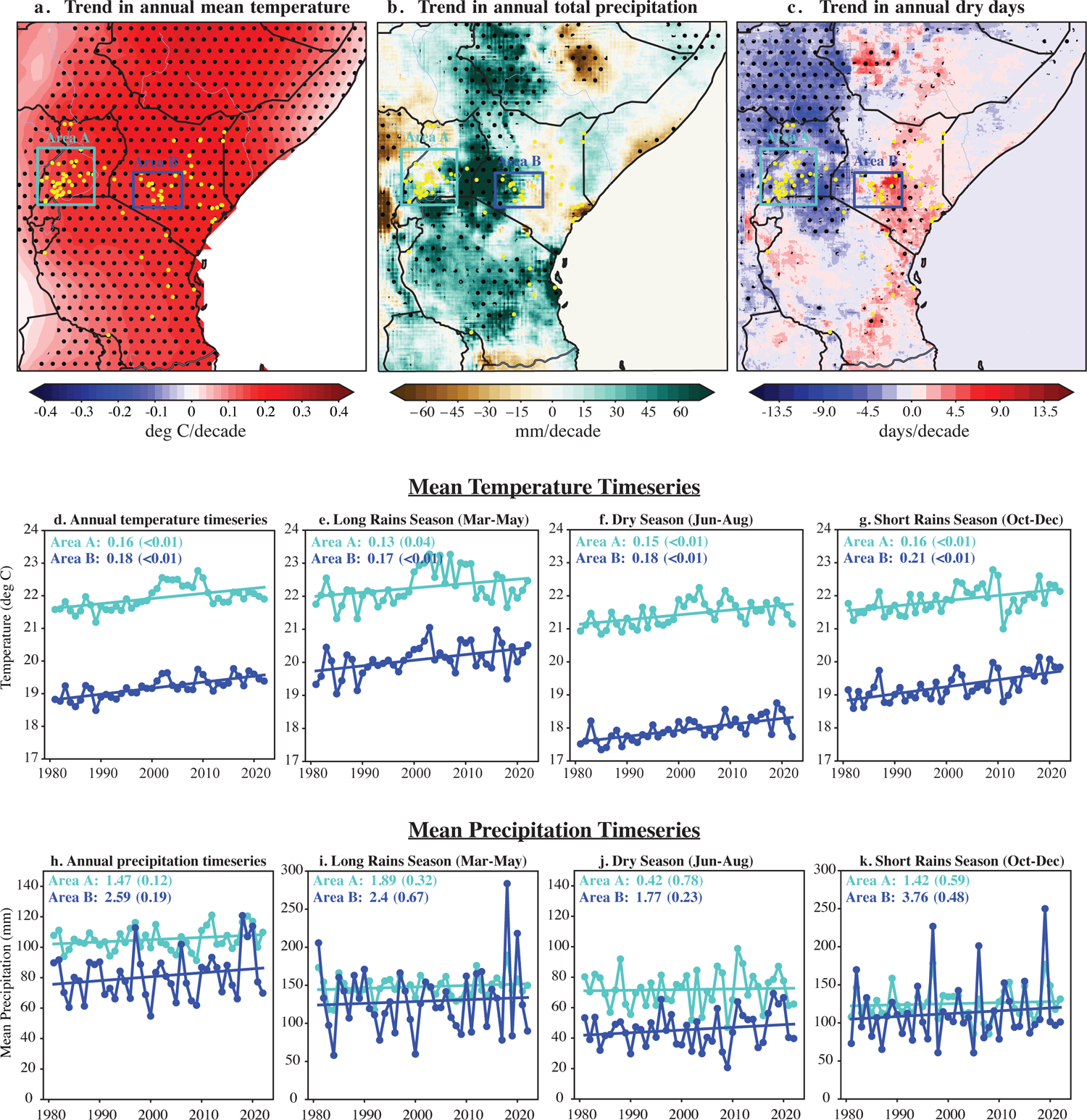
Effect of annual and seasonal temperature and rainfall on occurrence of RVF incidences

(Figure 3a,c) Long-term climatology (average over 1991-2020) and (Figure 3b,d) decadal trends (calculated from 1981-2022) of mean temperature during the two main rainy seasons - long rains (March-May) and short rains (October-December) season over East Africa. (Figure 3e-h) same as above but for total precipitation. Black dots in panels (Figure 3b,d,f) and (Figure 3h) represent areas with trends that are significant at the 10% level based on the Wald test.

Annual precipitation trends reflected the patterns of trends in seasonal rainfall, showing increased wetness across southern and eastern Uganda, western Kenya, and Tanzania (Figure 4b). Another important climate variability parameter is trends in annual number of dry days (<1mm of rain/day).

There has been a decreasing trend in the number of dry days (i.e., trend towards more rainy days) across most of Uganda, while the number of dry days is increasing across much of Kenya and parts of Tanzania (Figure 4c). Parts of Kenya with a high number of clusters experienced dry day increases of ∼3-6 days/decade while parts of Uganda with a high number of clusters experienced dry day decreases of 3-4.5 days/decade, despite increasing trends in annual rainfall (Figure 4b-c).

### Long-term climate variability in areas with a high density of RVF clusters

Because of limited number of RVF clusters from Tanzania, we use the data from Kenya and Uganda to assess the impact of climatic variability on incidence of RVF clusters. The assessment compared annual and seasonal mean temperature and rainfall trends in two regions; Area A in southwestern Uganda here 76% of the RVF clusters of the country occurred (Figure 4a-tourquoise box), and Area B in southwestern Kenya where 30% of in-country clusters occurred (Figure 4a-blue box). Timeseries of temperature and total rainfall during the long-rains (March-May), intervening dry season (June-August), and short-rains (October-December) in the RVF cluster Area A and Area B were compared (Figure 4d-k).

The two areas with a high concentration of RVF cluster areas (Figure 4a, Area A & Area B) in Uganda and Kenya experienced significant warming trends (p<0.05) across all seasons and annually over the past four decades. Area B in Kenya experienced slightly stronger warming than in Area A. (Figure 4d-g). For example, during the short-rain season, temperature in Area A increased at 0.16°C/decade while in Area B temperature increased at 0.21 °C/decade (Figure 4g). Relative to other areas in Uganda, parts of Area A experienced weaker annual warming while annual trends in Area B were similar in magnitude to trends across other parts of Kenya (Figure 4a). The consistent 2-3°C higher temperature in Uganda across all seasons (Figure 4d-g) is not unique to high concentration RVF cluster sites but rather reflects the normal climatological difference across the two regions.

Annual total rainfall over the high concentrations RVF cluster areas showed increasing trends in Area B in Kenya being approximately 1.7 times higher than in Area A (Figure 4h), although trends are not statistically significant. Similarly, although not significant, rainfall over the Area B in Kenya showed a faster increasing trend relative to Area A during the dry season and short rains season but not in the long-rains season (Figure 41-k). Trends in the short-rain season in Area B were >2.5 times higher than in Area A.

Top panels decadal trends (calculated from 1981-2022) in (Figure 4a) annual temperature, (Figure 4b) annual rainfall, and (Figure 4c) annual number of dry days (days with <1 mm/day) overlayed with the RVF clusters (yellow dots) in Kenya, Uganda, and Tanzania. Black dots in (Figure 4a-c) represent trends that are significant at the 10% level based on the Wald test. The turquoise and blue boxes in panel (Figure 4a) indicates RVF clusters in Uganda and Kenya, respectively. Timeseries of area-average, mean temperature (Figure 4d-g) and rainfall (Figure 4h-k) in the RVF cluster Area A (Uganda) and Area B (Kenya) calculated annual and for the long-rains, dry and short-rains seasons. Numbers in each panel represent the decadal trend magnitude (in deg C/decade in panels d-g and mm/decade in (Figure 4h-k) and p-values of the linear trend in brackets for the corresponding timeseries.

A timeseries plots of number of RVF clusters in the two hotspots; Area A in Uganda (Figure 5-top panel) and Area B in Kenya (Figure 5-bottom panel) during the long-rain seasons showed increasing trend as temperature and rainfall increased in these highland regions.

**Figure 5.**
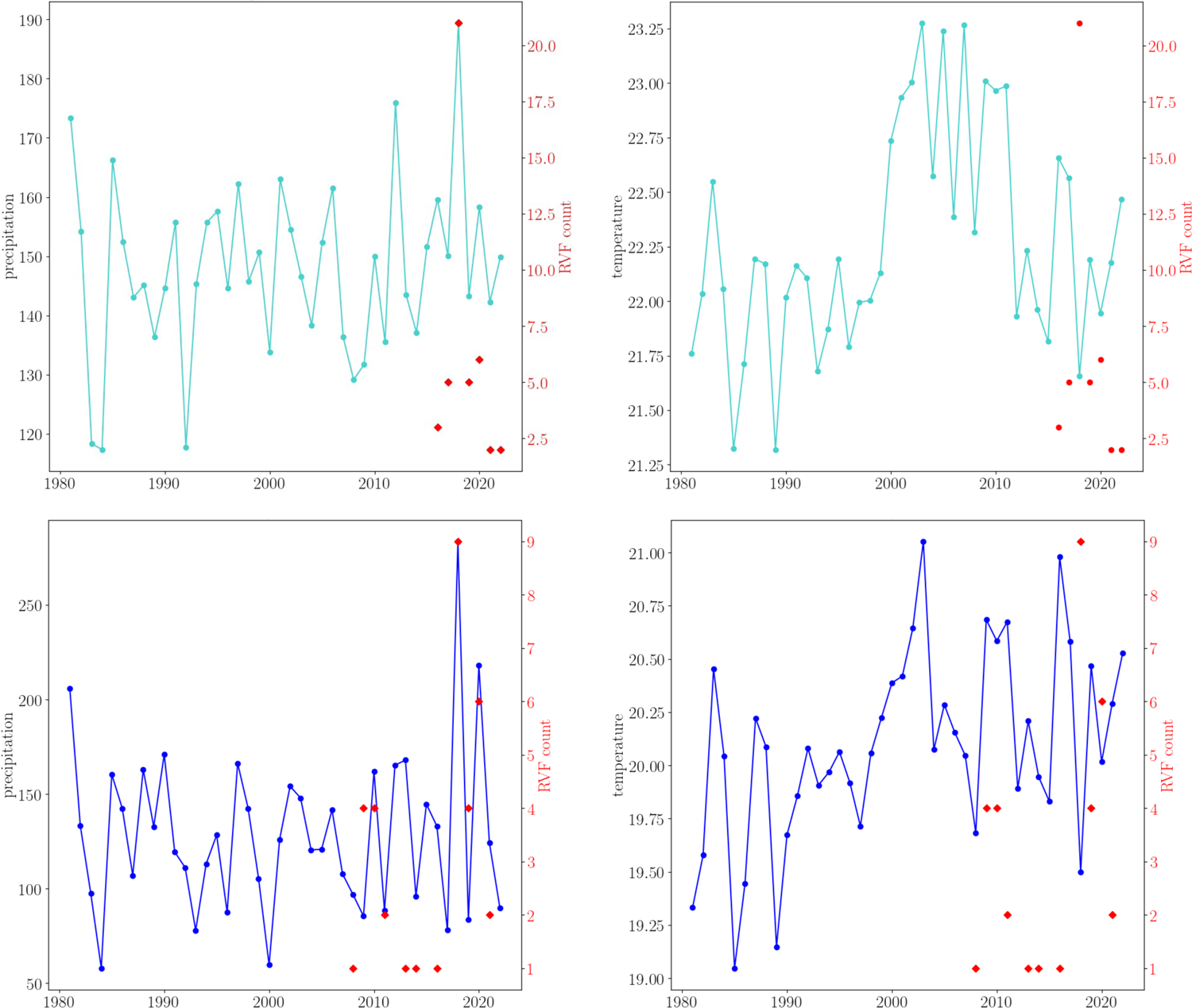
Increasing frequency of RVF clusters in two highland area of Kenya and Uganda associated with increasing precipitation and temperature.

## DISCUSSION

We performed spatiotemporal modeling involving various geo-ecological and short-term climatic factors associated with the increasing occurrence of small RVF clusters reported across widening geographic regions in East Africa in recent years. In addition, we analyzed long-term temperature and precipitation metrics trends in the region over four decades (1981-2022) and evaluated the association of these trends with hotspots of RVF clusters in Kenya and Uganda. The described RVF clusters are characterized by <5 human cases and <10 livestock cases. Notably, 35% of these clusters occurred in geographic areas that had not previously reported RVF disease. The small RVF clusters occurred during or immediately after rainy seasons, highlighting the critical role that mosquito vectors continue to play in the dissemination of the RVF virus. Comparable trends of small RVF clusters have been documented in various African countries in recent years. [53] [54] In contrast to the large epidemics associated with extreme weather and primarily occurring in areas of low elevation, specific soil types, and extensive livestock production, the small RVF clusters occurred in diverse ecosystems. These included both highlands and lowlands with varied soil types and encompassing the entire spectrum of livestock production systems; (intensive, semi-intensive, and extensive). Notably, a substantial proportion of clusters in Uganda (85%) and Kenya (55%) occurred in the highlands where mixed farming with intensive and semi-intensive livestock production was prevalent.

When we evaluated >40 independent variables for association with RVF occurrence, comparable short-term and long-term climatic factors emerged as primary drivers shaping the evolving landscape of RVF disease. Warmer temperatures and rainfall 1-3 months prior correlated with a higher RVF incidence rate ratio, with 80% of the clusters occurring during this phase. Factors crucial for extreme weather associated RVF epidemics like excessive flooding, low elevation, clayey soil types and extensive livestock production systems were not significant for small RVF cluster occurrence. Our regional analysis of long-term climatic trends revealed an annual mean temperature increase of up to 0.12° to 0.3°C per decade in parts of East Africa, indicating a rise in annual average temperatures of up to 1.2°C over the past 40 years. These climatic variations have directly contributed to escalating frequency and severity of droughts, particularly in the arid and semi-arid lowlands such as northeastern Kenya. [55] [56] Our findings on climatic trends analysis reveal increasing rainfall trends in scattered highlands of the regions. Importantly, our data indicates that areas in the highlands of southwestern Uganda and Kenya experiencing rising temperature and rainfall trends alongside diverging trends in dry days coincide with significantly increasing frequency of RVF clusters. The disease landscape in Uganda more vividly reflects the impact of climatic variations. Despite not being part of regional RVF epidemics and having no documented significant human RVF clusters for decades, the country has reported over 60 small RVF clusters across the country since 2016.

Our findings suggest that the combination of increasing temperature, increasing rainfall trends; and a decrease in the annual number of dry days is a driving factor behind the escalating public health risk of RVF disease in Uganda. With ongoing global warming, East Africa is projected to undergo changes in temperature and precipitation patterns with continued increases in seasonal and annual mean temperatures, particularly in highland areas. Seasonal and annual mean temperatures are expected to keep rising with the most significant increases occurring in the highland areas. [57] Even with a 2°C of global warming aligning with the Paris climate Agreement goals, annual temperatures in the region may increase by 1.5-2°C accompanied by over 40% increase in the number of days with temperature above 35°C. [58] In terms of precipitation, the observed wetting during the short rain season is projected to persist, accompanied by an increase in heavy rain event whereas the changes in rainfall patterns during the long rain season remain uncertain. [59] [60] The likely impact of these future climatic trends is a heightened frequency of small RVF clusters across the geographic areas described in our study, with a reduced frequency of occurrence for the extreme weather-associated large epidemics in the region’s lowlands.

The global impact of the spreading RVF clusters includes a wider dispersal of virus leading to the creation of new regions with virus endemicity. This escalation increases the risk of more widespread epidemics or pandemics in the future. Changing climate conditions and drivers of natural climate variability such as El Niño Southern Oscillation that brings extensive heavy rains and flooding across Africa and other regions can trigger widespread virus amplification and human infections. [60] [61] This is particularly concerning in rural Africa and Middle East where there is significant populations of livestock to support virus amplification and enhanced human infections through consumption of animal products. [60] [61].Studies have shown widespread distribution of *Aedes* mosquito vectors globally, including Europe, Australia and North America. [62] It is crucial for global public health institutions to consistently approve human RVF vaccines therapeutics and implement swift distribution plans through regional banks. [63] [64]

Fortunately, the highly conserved RVF virus genome and minimal antigenic variation observed during epidemics guarantees the high efficacy of vaccines and immunotherapeutics. [65] [66] The study has limitations. First, reliance on available data introduces potential bias because of the incompleteness of some of the data points and underreporting in regions with limited health infrastructure. In addition, the presence of potential multicollinearity of variables and the use of presence-only data should be considered when interpreting the data. Conversely, an increase in disease reporting rate over time and in new geographic areas is difficult to disentangle from increases in actual disease prevalence.

While we acknowledge these potential limitations, our analyses suggest distinct ecological and climatic risk factors underlying the changing epidemiology of RVF disease in East Africa, which should be investigated forin direct causal relationship. Specifically, our study highlights the increasing frequency of small RVF clusters in previously unaffected areas, associated with a combination of higher temperature and rainfall.

## Supporting information

Supplemental Table 1.Predictor variables of Rift Valley Fever disease cluster occurrence analyzed.

## Data Availability

The data used to support the findings of this study can be provided from the authors on request.

## Acknowledgment

We acknowledge the collaborative support from the Ministries of Health and Agriculture in Kenya and Uganda. Additionally, we express our gratitude to the Kenya Medical Research Institute (KEMRI) and the Directorate of Veterinary Services for their essential contributions to this research.

## Author contributions

S.S., L.N, M.K.N, and D.S; conceptualization, data curation, formal analysis, writing original draft and writing-review and editing; I.N.; E.O.; E.C;E.L; E.O, and S.O.O; methodology, formal analysis, supervision, and writing-review and editing M.M.; M.M.; M.M; A.M.; L.K.; S.K.; I.N.; R.B.; B.B.; A.M.; supervision, writing-review and editing, J.G. data curation, methodology, formal analysis, supervision and writing-review and editing; J.D.; J.G.; D.S.; M.K.N.; conceptualization, data curation, methodology, formal analysis, supervision and writing-review and editing; conceptualization, methodology, formal analysis, supervision and writing-review and editing. All authors gave final approval for publication and agreed to be held accountable for the work performed therein.

## Patient and public involvement

A policy statement summarizing the finding and implication of these results was developed and disseminated to Ministries of Health of Kenya, Uganda, and Tanzania.

## REFERENCES

[1] D. Wright, J. Kortekaas, T. A. Bowden, and G. M. Warimwe, “Rift Valley fever: biology and epidemiology,” Journal of General Virology, vol. 100, no. 8, pp. 1187–1199, Aug. 2019, doi: 10.1099/jgv.0.001296.

[2] M. Pepin, M. Bouloy, B. H. Bird, A. Kemp, and J. Paweska, “Rift Valley fever virus: an update on pathogenesis, molecular epidemiology, vectors, diagnostics and prevention,” Vet. Res., vol. 41, no. 6, p. 61, Nov. 2010, doi: 10.1051/vetres/2010033.

[3] R. M. Murithi et al., “Rift Valley fever in Kenya: history of epizootics and identification of vulnerable districts,” Epidemiol. Infect., vol. 139, no. 3, pp. 372–380, Mar. 2011, doi: 10.1017/S0950268810001020.

[4] P. M. Nguku et al., “An investigation of a major outbreak of Rift Valley fever in Kenya: 2006-2007,” Am J Trop Med Hyg, vol. 83, no. 2 Suppl, pp. 5–13, Aug. 2010, doi: 10.4269/ajtmh.2010.09-0288.

[5] K. Ahmad, “More deaths from Rift Valley fever in Saudi Arabia and Yemen,” The Lancet, vol. 356, no. 9239, p. 1422, Oct. 2000, doi: 10.1016/S0140-6736(05)74068-X.

[6] Centers for Disease Control and Prevention (CDC), “Outbreak of Rift Valley fever--Saudi Arabia, August-October, 2000,” MMWR Morb Mortal Wkly Rep, vol. 49, no. 40, pp. 905–908, Oct. 2000.

[7] R. Sang et al., “Rift Valley Fever Virus Epidemic in Kenya, 2006/2007: The Entomologic Investigations,” The American Journal of Tropical Medicine and Hygiene, vol. 83, no. 2_Suppl, pp. 28–37, Aug. 2010, doi: 10.4269/ajtmh.2010.09-0319.

[8] A. Hightower et al., “Relationship of Climate, Geography, and Geology to the Incidence of Rift Valley Fever in Kenya during the 2006–2007 Outbreak,” The American Journal of Tropical Medicine and Hygiene, vol. 86, no. 2, pp. 373–380, Feb. 2012, doi: 10.4269/ajtmh.2012.11-0450.

[9] A. Mohamed, H. Ghazi, A. Ashshi, H. Faidah, and E. Azhar, “Serological Survey of Rift Valley Fever among Sacrifice Animals in Holy Mecca During Pilgrimage Season,” International Journal of Tropical Medicine, vol. 6, pp. 85–89, Jan. 2011.

[10] Y. E. Himeidan, E. J. Kweka, M. M. Mahgoub, E. A. El Rayah, and J. O. Ouma, “Recent Outbreaks of Rift Valley Fever in East Africa and the Middle East,” Front. Public Health, vol. 2, Oct. 2014, doi: 10.3389/fpubh.2014.00169.

[11] F. G. Davies, K. J. Linthicum, and A. D. James, “Rainfall and epizootic Rift Valley fever,” Bull World Health Organ, vol. 63, no. 5, pp. 941–943, 1985.

[12] K. J. Linthicum, F. G. Davies, A. Kairo, and C. L. Bailey, “Rift Valley fever virus (family Bunyaviridae, genus Phlebovirus). Isolations from Diptera collected during an inter-epizootic period in Kenya,” J Hyg (Lond), vol. 95, no. 1, pp. 197–209, Aug. 1985, doi: 10.1017/s0022172400062434.

[13] M. Kariuki Njenga and B. Bett, “Rift Valley Fever Virus—How and Where Virus Is Maintained During Inter-epidemic Periods,” Curr Clin Micro Rpt, vol. 6, no. 1, pp. 18–24, Mar. 2019, doi: 10.1007/s40588-018-0110-1.

[14] A. Evans et al., “Prevalence of antibodies against Rift Valley fever virus in Kenyan wildlife,” Epidemiol. Infect., vol. 136, no. 9, pp. 1261–1269, Sep. 2008, doi: 10.1017/S0950268807009806.

[15] M. J. Turell, A. R. Spring, M. K. Miller, and C. E. Cannon, “Effect of Holding Conditions on the Detection of West Nile Viral RNA by Reverse Transcriptase-Polymerase Chain Reaction from Mosquito (Diptera: Culicidae) Pools: Table 1,” J Med Entomol, vol. 39, no. 1, pp. 1–3, Jan. 2002, doi: 10.1603/0022-2585-39.1.1.

[16] M. J. Turell, K. J. Linthicum, L. A. Patrican, F. G. Davies, A. Kairo, and C. L. Bailey, “Vector Competence of Selected African Mosquito (Diptera: Culicidae) Species for Rift Valley Fever Virus,” J Med Entomol, vol. 45, no. 1, pp. 102–108, Jan. 2008, doi: 10.1093/jmedent/45.1.102.

[17] E. A. Gould and S. Higgs, “Impact of climate change and other factors on emerging arbovirus diseases,” Transactions of the Royal Society of Tropical Medicine and Hygiene, vol. 103, no. 2, pp. 109–121, Feb. 2009, doi: 10.1016/j.trstmh.2008.07.025.

[18] K. J. Linthicum, S. C. Britch, and A. Anyamba, “Rift Valley Fever: An Emerging Mosquito-Borne Disease,” Annu Rev Entomol, vol. 61, pp. 395–415, 2016, doi: 10.1146/annurev-ento-010715-023819.

[19] T. R. Shoemaker et al., “First Laboratory-Confirmed Outbreak of Human and Animal Rift Valley Fever Virus in Uganda in 48 Years,” The American Journal of Tropical Medicine and Hygiene, vol. 100, no. 3, pp. 659–671, Mar. 2019, doi: 10.4269/ajtmh.18-0732.

[20] D. Tumusiime et al., “Mapping the risk of Rift Valley fever in Uganda using national seroprevalence data from cattle, sheep and goats,” PLoS Negl Trop Dis, vol. 17, no. 5, p. e0010482, May 2023, doi: 10.1371/journal.pntd.0010482.

[21] World Food Program, “Eastern Africa seasonal monitor,” 2022. [Online]. Available: https://docs.wfp.org/api/documents/WFP-0000142577/download/

[22] C. Sindato, K. B. Stevens, E. D. Karimuribo, L. E. G. Mboera, J. T. Paweska, and D. U. Pfeiffer, “Spatial Heterogeneity of Habitat Suitability for Rift Valley Fever Occurrence in Tanzania: An Ecological Niche Modelling Approach,” PLoS Negl Trop Dis, vol. 10, no. 9, p. e0005002, Sep. 2016, doi: 10.1371/journal.pntd.0005002.

[23] R. D. Sumaye, E. Geubbels, E. Mbeyela, and D. Berkvens, “Inter-epidemic Transmission of Rift Valley Fever in Livestock in the Kilombero River Valley, Tanzania: A Cross-Sectional Survey,” PLoS Negl Trop Dis, vol. 7, no. 8, p. e2356, Aug. 2013, doi: 10.1371/journal.pntd.0002356.

[24] E. G. Kifaro et al., “Epidemiological study of Rift Valley fever virus in Kigoma, Tanzania,” Onderstepoort J Vet Res, vol. 81, no. 2, p. 5 pages, Apr. 2014, doi: 10.4102/ojvr.v81i2.717.

[25] R. D. Sumaye, E. N. Abatih, E. Thiry, M. Amuri, D. Berkvens, and E. Geubbels, “Inter-epidemic Acquisition of Rift Valley Fever Virus in Humans in Tanzania,” PLoS Negl Trop Dis, vol. 9, no. 2, p. e0003536, Feb. 2015, doi: 10.1371/journal.pntd.0003536.

[26] J. J. Wensman et al., “A study of Rift Valley fever virus in Morogoro and Arusha regions of Tanzania – serology and farmers’ perceptions,” Infection Ecology & Epidemiology, vol. 5, no. 1, p. 30025, Jan. 2015, doi: 10.3402/iee.v5.30025.

[27] WHO, “Weekly bulletin on outbreaks and other emergencies,” WHO, 2017. [Online]. Available: https://apps.who.int/iris/bitstream/handle/10665/255895/OEW28-81472017.pdf;jsessionid=75C9C66D6CAF054F1D2BE7687D5BB253?sequence=1

[28] A. Ahmed, J. Makame, F. Robert, K. Julius, and M. Mecky, “Sero-prevalence and spatial distribution of Rift Valley fever infection among agro-pastoral and pastoral communities during Interepidemic period in the Serengeti ecosystem, northern Tanzania,” BMC Infect Dis, vol. 18, no. 1, p. 276, Dec. 2018, doi: 10.1186/s12879-018-3183-9.

[29] M. K. Matiko, L. P. Salekwa, C. J. Kasanga, S. I. Kimera, M. Evander, and W. P. Nyangi, “Serological evidence of inter-epizootic/inter-epidemic circulation of Rift Valley fever virus in domestic cattle in Kyela and Morogoro, Tanzania,” PLoS Neglected Tropical Diseases, vol. 12, no. 11, Nov. 2018, doi: 10.1371/journal.pntd.0006931.

[30] L. Nyakarahuka et al., “Ten outbreaks of rift valley fever in Uganda 2016-2018: epidemiological and laboratory findings,” International Journal of Infectious Diseases, vol. 79, p. 4, Feb. 2019, doi: 10.1016/j.ijid.2018.11.029.

[31] L. P. Salekwa, P. N. Wambura, M. K. Matiko, and D. M. Watts, “Circulation of Rift Valley Fever Virus Antibody in Cattle during Inter-Epizootic/Epidemic Periods in Selected Regions of Tanzania,” The American Journal of Tropical Medicine and Hygiene, vol. 101, no. 2, pp. 459–466, Aug. 2019, doi: 10.4269/ajtmh.18-0798.

[32] R. M. Budodo, P. G. Horumpende, S. I. Mkumbaye, B. T. Mmbaga, R. S. Mwakapuja, and J. O. Chilongola, “Serological evidence of exposure to Rift Valley, Dengue and Chikungunya Viruses among agropastoral communities in Manyara and Morogoro regions in Tanzania: A community survey,” PLoS Negl Trop Dis, vol. 14, no. 7, p. e0008061, Jul. 2020, doi: 10.1371/journal.pntd.0008061.

[33] M. J. Nyarobi, “The epidemiology of Rift Valley fever in northern Tanzania.,” 2020. [Online]. Available: https://theses.gla.ac.uk/81309/

[34] S. Rugarabamu et al., “Seroprevalence and associated risk factors of selected zoonotic viral hemorrhagic fevers in Tanzania,” International Journal of Infectious Diseases, vol. 109, pp. 174–181, Aug. 2021, doi: 10.1016/j.ijid.2021.07.006.

[35] M. S. Kumalija et al., “Detection of Rift Valley Fever virus inter-epidemic activity in Kilimanjaro Region, North Eastern Tanzania,” Global Health Action, vol. 14, no. 1, p. 1957554, Jan. 2021, doi: 10.1080/16549716.2021.1957554.

[36] WHO, “Disease Outbreak News,” WHO. Accessed: Jul. 19, 2023. [Online]. Available: https://www.who.int/emergencies/disease-outbreak-news

[37] WAHIS, “World Organisation for Animal Health (WOAH, founded as OIE).,” WOAH - World Organisation for Animal Health. Accessed: Jul. 19, 2023. [Online]. Available: https://www.woah.org/en/home/

[38] W. A. De Glanville et al., “An outbreak of Rift Valley fever among peri-urban dairy cattle in northern Tanzania, 2018,” Ecology, preprint, Sep. 2021. doi: 10.1101/2021.09.15.459147.

[39] MODIS, “MODIS.” Accessed: Jul. 19, 2023. [Online]. Available: https://modis.gsfc.nasa.gov/data/

[40] FAO, “Relative humidity at 12h local time - AgERA5 (Global - Daily - ∼10km) - ECMWF - European Centre for Medium-Range Weather Forecasts - ‘FAO catalog.’” Accessed: Oct. 05, 2023. [Online]. Available: https://data.apps.fao.org/catalog/dataset/7cb8355c-e935-406b-bd02-f48f0ed4543c/resource/f997f268-585c-4017-a9a0-cc4207367797

[41] Kenya National Bureau of Statistics, “2019 Kenya Population and Housing Census Results - Kenya National Bureau of Statistics.” Accessed: Jul. 19, 2023. [Online]. Available: https://www.knbs.or.ke/2019-kenya-population-and-housing-census-results/

[42] Uganda Bureau of statistics, “National population and housing census,” 2014. [Online]. Available: https://unstats.un.org/unsd/demographic-social/census/documents/Uganda/UGA-2014-11.pdf

[43] Tanzania census Information dissemination platform, “Census Information dissemination platform.” Accessed: Jul. 19, 2023. [Online]. Available: https://sensa.nbs.go.tz/publications

[44] Tanzania National Bureau of statistics, “National Bureau of statistics,” 2012. [Online]. Available: https://www.ilo.org/ilostat-files/SSM/SSM9/TANZANIA.pdf

[45] United Nations population Fund, “World Population Dashboard.” Accessed: Jul. 19, 2023. [Online]. Available: https://www.unfpa.org/data/world-population-dashboard

[46] CHIRPS, “Rainfall Data Sets.” Accessed: Jul. 15, 2023. [Online]. Available: https://docs.digitalearthafrica.org/en/latest/data_specs/CHIRPS_specs.html#:~:text=Climate%20Hazards%20Group%20InfraRed%20Precipitation%20with%20Station%20data%20(CHIRPS)%20is,from%201981%20to%20near%2Dpresent.https://www.watres.com/CHIRPS/

[47] CHIRTS, “CHIRTS | Climate Hazards Center,” CHIRTSdaily | Climate Hazards Center - UC Santa Barbara. Accessed: Oct. 05, 2023. [Online]. Available: https://www.chc.ucsb.edu/data/chirtsdaily

[48] C. Funk et al., “The climate hazards infrared precipitation with stations—a new environmental record for monitoring extremes,” Sci Data, vol. 2, no. 1, p. 150066, Dec. 2015, doi: 10.1038/sdata.2015.66.

[49] C. Funk et al., “A High-Resolution 1983–2016 Tmax Climate Data Record Based on Infrared Temperatures and Stations by the Climate Hazard Center,” Journal of Climate, vol. 32, no. 17, pp. 5639–5658, Sep. 2019, doi: 10.1175/JCLI-D-18-0698.1.

[50] G. Lo Iacono, A. A. Cunningham, B. Bett, D. Grace, D. W. Redding, and J. L. N. Wood, “Environmental limits of Rift Valley fever revealed using ecoepidemiological mechanistic models,” Proc Natl Acad Sci U S A, vol. 115, no. 31, pp. E7448–E7456, Jul. 2018, doi: 10.1073/pnas.1803264115.

[51] G. Mosomtai et al., “Association of ecological factors with Rift Valley fever occurrence and mapping of risk zones in Kenya,” Int J Infect Dis, vol. 46, pp. 49–55, May 2016, doi: 10.1016/j.ijid.2016.03.013.

[52] J. C. Jakobsen, C. Gluud, J. Wetterslev, and P. Winkel, “When and how should multiple imputation be used for handling missing data in randomised clinical trials – a practical guide with flowcharts,” BMC Medical Research Methodology, vol. 17, no. 1, p. 162, Dec. 2017, doi: 10.1186/s12874-017-0442-1.

[53] P. Jansen van Vuren et al., “Human Cases of Rift Valley Fever in South Africa, 2018,” Vector-Borne and Zoonotic Diseases, vol. 18, no. 12, pp. 713–715, Dec. 2018, doi: 10.1089/vbz.2018.2357.

[54] D. Sissoko et al., “Rift Valley Fever, Mayotte, 2007–2008,” Emerg. Infect. Dis., vol. 15, no. 4, pp. 568–570, Apr. 2009, doi: 10.3201/eid1504.081045.

[55] UNEP, “UNEP Climate Action Note | Data you need to know.” Accessed: Sep. 21, 2023. [Online]. Available: https://www.unep.org/explore-topics/climate-action/what-we-do/climate-action-note/state-of-the-climate.html

[56] M. Kilavi et al., “Extreme Rainfall and Flooding over Central Kenya Including Nairobi City during the Long-Rains Season 2018: Causes, Predictability, and Potential for Early Warning and Actions,” Atmosphere, vol. 9, no. 12, p. 472, Nov. 2018, doi: 10.3390/atmos9120472.

[57] C. H. Trisos et al., “Africa. In: Climate Change 2022: Impacts, Adaptation and Vulnerability. Contribution of Working Group II to the Sixth Assessment Report of the Intergovernmental Panel on Climate Change [H.-O. Pörtner, D.C. Roberts, M. Tignor, E.S. Poloczanska, K. Mintenbeck, A. Alegría, M. Craig, S. Langsdorf, S. Löschke, V. Möller, A. Okem, B. Rama (eds.)],” Cambridge University Press, Cambridge, UK and New York, NY, USA, 2022. [Online]. Available: doi:10.1017/9781009325844.011.

[58] J. M. Gutiérrez et al., “The Physical Science Basis. Contribution of Working Group I to the Sixth Assessment Report of the Intergovernmental Panel on Climate Change [Masson-Delmotte, V., P. Zhai, A. Pirani, S.L. Connors, C. Péan, S. Berger, N. Caud, Y. Chen, L. Goldfarb, M.I. Gomis, M. Huang, K. Leitzell, E. Lonnoy, J.B.R. Matthews, T.K. Maycock, T. Waterfield, O. Yelekçi, R. Yu, and B. Zhou (eds.)].,” Cambridge University Press, United Kingdom and New York, NY, USA, 2021. [Online]. Available: doi:10.1017/9781009157896.021

[59] P. I. Palmer et al., “Drivers and impacts of Eastern African rainfall variability,” Nat Rev Earth Environ, vol. 4, no. 4, pp. 254–270, Mar. 2023, doi: 10.1038/s43017-023-00397-x.

[60] S. Amwayi et al., “Risk factors for severe rift valley fever infection in Kenya, 2007.” Accessed: Sep. 21, 2023. [Online]. Available: https://pure.johnshopkins.edu/en/publications/risk-factors-for-severe-rift-valley-fever-infection-in-kenya-2007

[61] M. O. Nanyingi et al., “A systematic review of Rift Valley Fever epidemiology 1931–2014,” Infection Ecology & Epidemiology, vol. 5, no. 1, p. 28024, Jan. 2015, doi: 10.3402/iee.v5.28024.

[62] S. Leta, T. J. Beyene, E. M. De Clercq, K. Amenu, M. U. G. Kraemer, and C. W. Revie, “Global risk mapping for major diseases transmitted by Aedes aegypti and Aedes albopictus,” International Journal of Infectious Diseases, vol. 67, pp. 25–35, Feb. 2018, doi: 10.1016/j.ijid.2017.11.026.

[63] WHO, “Efficacy trials of Rift Valley Fever vaccines and therapeutics Guidance on clinical trial design,” WHO, R&D Blueprint, 2023.

[64] CEPI, “Rift Valley fever vaccines,” CEPI. Accessed: Sep. 21, 2023. [Online]. Available: https://cepi.net/news_cepi/rift-valley-fever-vaccines-to-advance-with-new-50-million-cepi-and-eu-funding-call/

[65] B. H. Bird, M. L. Khristova, P. E. Rollin, T. G. Ksiazek, and S. T. Nichol, “Complete Genome Analysis of 33 Ecologically and Biologically Diverse Rift Valley Fever Virus Strains Reveals Widespread Virus Movement and Low Genetic Diversity due to Recent Common Ancestry,” J Virol, vol. 81, no. 6, pp. 2805–2816, Mar. 2007, doi: 10.1128/JVI.02095-06.

[66] J. Juma et al., “Genomic surveillance of Rift Valley fever virus: from sequencing to lineage assignment,” BMC Genomics, vol. 23, no. 1, p. 520, Dec. 2022, doi: 10.1186/s12864-022-08764-6.

