## Supplemental Table 1.Predictor variables of Rift Valley Fever disease cluster occurrence analyzed. for "Widening geographic range of Rift Valley fever disease clusters associated with climate change in East Africa"

**Supplementary Table I.** Predictor variables of Rift Valley Fever disease cluster occurrence analyzed.

| | Variable | **Units** |
|---|---|---|
| 1 | Country | NA |
| 2 | Region | NA |
| 3 | County or District | NA |
| 4 | Year of outbreak | Year |
| 5 | Month of outbreak | Month |
| 6 | History of absence of previous outbreak | NA |
| 7 | History of previous outbreaks | NA |
| 8 | *Land factors* | |
| 9 | Soil type | NA |
| 10 | Altitude | Meters |
| 11 | Elevation below 1000 m | Meters |
| 12 | Presence of land use changes | NA |
| 13 | Types of land use changes | NA |
| 14 | Presence of significant water pools | NA |
| 15 | Causes of significant water pools | NA |
| 16 | *Animal factors* | |
| 17 | Predominant livestock species | NA |
| 18 | Type of Livestock production system | NA |
| 19 | Host density | Population/ $Km^2$ |
| 20 | Proximity to wildlife | Km |
| 21 | *Climate data* | |
| 22 | Annual rainfall 3 years prior to the outbreak | mm |
| 23 | Annual rainfall 2 years prior to the outbreak | mm |
| 24 | Annual rainfall for the year preceding the outbreak | mm |
| 25 | Annual rainfall of the outbreak year | mm |
| 26 | Monthly rainfall for the outbreak month | mm |
| 27 | Monthly rainfall 1 month prior to the outbreak | mm |
| 28 | Monthly rainfall 2 months prior to the outbreak | mm |
| 29 | Monthly rainfall 3 months prior to the outbreak | mm |
| 30 | Annual temperature 3 years prior to the outbreak | °Celsius |
| 31 | Annual temperature 2 years prior to the outbreak | °Celsius |
| 32 | Annual temperature for the year preceding the outbreak | °Celsius |
| 33 | Annual temperature of the outbreak year | °Celsius |
| 34 | Monthly temperature for the outbreak month | °Celsius |
| 35 | Monthly temperature 1 month prior to the outbreak | °Celsius |
| 36 | Monthly temperature 2 months prior to the outbreak | °Celsius |
| 37 | Monthly temperature 3 months prior to the outbreak | °Celsius |
| 38 | Annual humidity 3 years prior to the outbreak | % |
| 39 | Annual humidity 2 years prior to the outbreak | % |
| 40 | Annual humidity for the year preceding the outbreak | % |
| 41 | Annual humidity for the year of the outbreak | % |

| | | |
|---|---|---|
| 42 | Monthly humidity for the outbreak month | % |
| 43 | Monthly humidity 1 month prior to the outbreak | % |
| 44 | Monthly humidity 2 months prior to the outbreak | % |
| 45 | Monthly humidity 3 months prior to the outbreak | % |
| 46 | Outbreak month NDVI | NA |
| 47 | Monthly NDVI 1 month prior to the outbreak | NA |
| 48 | Monthly NDVI 2 months prior to the outbreak | NA |
| 49 | Monthly NDVI 3 months prior to the outbreak | NA |
