## Supplemental Data 1 for "Widening geographic range of Rift Valley fever disease clusters associated with climate change in East Africa"

**Supplement Table II.** Univariate analysis of drivers of RVF clusters in East Africa, 2008 –2022

| Variable | Coefficient | IRR | 95% CI | S.E | p-value |
|---|---|---|---|---|---|
| Country* | | | | | |
| *Kenya (Intercept)* | -3.86 | - | 0.01, 0.03 | 0.19 | <0.001 |
| *Uganda* | -1.55 | 0.21 | 0.12, 0.38 | 0.28 | <0.001 |
| *Tanzania* | 0.90 | 2.46 | 1.14, 5.66 | 0.39 | 0.02* |
| Livestock production system* | | | | | |
| *Extensive (Intercept)* | -3.41 | - | 0.02, 0.05 | 0.22 | <0.001 |
| *Intensive* | -0.63 | 0.531 | 1.13, 3.93 | 0.35 | 0.06 |
| *Semi-intensive* | -1.89 | 0.150 | 1.08, 1.32 | 0.31 | <0.001 |
| Elevation level* | | | | | |
| *High (> 1000m) (Intercept)* | -5.17 | - | 0.004, 0.008 | 0.15 | <0.001 |
| *Low (<500m)* | 2.29 | 9.96 | 5.81, 17.60 | 0.27 | <0.001 |
| *Medium (500 - 1000m)* | 0.47 | 1.61 | 0.68, 4.15 | 0.44 | 0.28 |
| Rainfall prior to outbreak (mm) | | | | | |
| 3$^{rd}$ month | 0.22 | 1.25 | 1.16, 1.34 | 0.0385 | <0.001 |
| 2$^{nd}$ month | 0.22 | **1.24** | 1.15, 1.34 | 0.0411 | <0.001 |
| 1$^{st}$ month | 0.18 | **1.19** | 1.11, 1.30 | 0.0458 | <0.001 |
| Outbreak month | 0.21 | **1.2438** | 1.13, 1.36 | 0.0523 | <0.001 |
| Humidity outbreak month | 0.04 | **1.04** | 1.01, 1.08 | 0.01751 | 0.01* |
| Positive change in NDVI (1$^{st}$month prior) | | | | | |
| *No (Intercept)* | - | - | | - | - |
| *Yes* | 0.31 | **1.36** | 0.71, 2.60 | 0.31 | 0.32 |
| NDVI change between 2$^{nd}$ month prior & outbreak month* | | | | | |
| *No (Intercept)* | - | - | | - | - |
| *Yes* | 4.60 | **99.74** | 13.01, 812.40 | 0.97 | <0.001 |

*Note S.E= standard error, IRR = incident rate, significant variables are in bold*
