## Supplemental Data 2 for "Widening geographic range of Rift Valley fever disease clusters associated with climate change in East Africa"

**Supplement Table III.** Multivariate negative binomial regression analysis of RVF cluster drivers in East Africa, 2008 –2022

| Factor | Coefficient | IRR | 95% CI | S.E | p-value |
|---|---|---|---|---|---|
| *Intercept* | -11.15 | - | - | 1.39 | <0.001 |
| Tanzania | 0.79 | 2.21 | 1.01, 5.20 | 0.39 | **0.04 *** |
| Uganda | -1.22 | 0.29 | 0.16, 0.53 | 0.29 | **<0.001** |
| Temperature (3rd month prior) | 0.18 | 1.20 | 1.11, 1.29 | 0.04 | **<0.001** |
| Positive NDVI change (1 month prior) | 0.58 | 1.79 | 1.00, 3.25 | 0.28 | **0.03 *** |
| First month prior NDVI | 1.59 | 4.9 | 0.93, 26.52 | 0.81 | **0.04 *** |
| Deviance | - | - | DF | - | - |
| Null Deviance | 161.50 | | 83 | - | - |
| Residual Deviance | 83.69 | | 78 | - | - |
| Dispersion parameter | 1.03 | | - | - | - |
| AIC | 449.55 | | - | - | - |
| 2 x log-likelihood | -435.55 | | - | - | - |

*Theta: 1.028; Std. Err.: 0.174, 0 '***' 0.001 '**' 0.01 '*' 0.05 '.' 0.1 ' ' 1*
